# Large-Language-Model Mortality Risk Stratification in the Intensive Care Unit: A Benchmark Against APACHE II

**DOI:** 10.1101/2025.05.14.25327650

**Authors:** Ahad Khaleghi Ardabili, Alireza Vafaei Sadr, Vida Abedi, Anthony S Bonavia

**Affiliations:** Department of Anesthesiology and Perioperative Medicine, Penn State Milton S Hershey Medical Center, Hershey, PA 17036, USA; Critical Illness and Sepsis Research Center (CISRC), Penn State College of Medicine, Hershey, PA 17036, USA; Department of Public Health Sciences, Penn State College of Medicine, Hershey, PA, USA; Division of Critical Care Medicine, Department of Anesthesiology and Perioperative Medicine, Penn State Milton S Hershey Medical Center, Hershey, PA 17036, USA

**Keywords:** large language models, sepsis, critical illness, prognostication, APACHE II

## Abstract

**Background:** Accurately predicting clinical trajectories in critically ill patients remains challenging due to physiological instability and multisystem organ dysfunction. Traditional prognostic tools, such as the APACHE II score, offer standardized risk assessment but are constrained by static algorithms. This study evaluates the predictive performance and reliability of large language models (LLMs) compared to APACHE II for in-hospital mortality prediction.

**Methods:** This was a single-center, retrospective study. De-identified clinical data from 70 critically ill patients were provided to four LLMs—Gemini, Llama, GPT-4, and R1. Each model stratified patients into high-, intermediate-, or low-risk (of in-hospital death) categories without being instructed to apply the APACHE II method. To assess the impact of additional information, models were also provided with de-identified hospital discharge summaries from prior hospital admissions. Consistency and rationale analyses were performed across multiple iterations.

**Findings:** LLMs demonstrated a general tendency toward risk overestimation, classifying more patients as high risk compared to APACHE II. Mortality rates within high-risk groups were lower than APACHE-predicted rates, suggesting calibration mismatch. Gemini, when supplemented with additional clinical context, uniquely identified a low-risk group. Gemini, GPT-4, and R1 exhibited the highest consistency across repeated evaluations, while Llama showed greater variability that improved with context. Semantic rationale analyses revealed greater stability among larger models, indicating non-stochastic reasoning patterns.

**Conclusions:** LLMs, supplemented with discharge summaries from prior hospitalizations, show promise in mortality risk stratification in critically ill patients. However, further refinement is necessary to improve calibration and reliability before clinical implementation. Context-aware prompting strategies and improved model calibration may enhance the utility of LLMs alongside established systems like APACHE II.

**Author Summary:** Predicting which critically ill patients are at greatest risk of dying in the hospital is one of the most important and difficult tasks faced by doctors. Traditionally, we’ve used structured scoring systems like APACHE II, which rely on a fixed set of patient measurements. In this study, we explored whether large language models (LLMs)—the same kind of technology behind chatbots like ChatGPT—could perform this task just as well, or even better. We provided four different LLMs with real patient data from our intensive care unit and asked them to assess each patient’s risk of dying, without giving them any instructions about how to do so. We also tested whether adding more context, such as hospital discharge summaries, made their predictions more accurate or consistent. We found that while LLMs tended to overestimate risk, some models—especially when given extra clinical information—showed strong consistency and thoughtful reasoning in their predictions. Our findings suggest that LLMs may eventually serve as helpful partners to physicians, offering a flexible and adaptable way to interpret complex clinical data. However, more work is needed to ensure that these tools are safe, reliable, and transparent before they can be used in real-world hospital settings

## Introduction

Accurately predicting in-hospital mortality in critically ill patients is vital for clinical decision-making but remains a persistent challenge. Established scoring systems like the Acute Physiology and Chronic Health Evaluation (APACHE) II offer standardized assessments of illness severity but are constrained by static variables and limited adaptability (1–3). Meanwhile, large language models (LLMs)—originally developed for natural language processing—have demonstrated potential in synthesizing complex clinical data and supporting medical decision-making (4–7).

Recent work has shown that LLMs can generate plausible clinical inferences (8–10), yet their reliability, consistency, and reasoning transparency in high-stakes contexts like critical care prognostication remain underexplored (11–13). Furthermore, their calibration and interpretability have not been rigorously compared to validated clinical tools.

This study benchmarks the performance of four LLMs—Gemini, GPT-4, R1, and Llama—against APACHE II in predicting in-hospital mortality using real-world intensive care unit data. We evaluate whether these models can produce consistent, interpretable, and clinically meaningful risk stratifications. Importantly, we examine whether providing additional clinical context (e.g., discharge summaries from prior hospitalizations) improves their predictions. By focusing on accuracy, consistency, and rationale stability, this study aims to inform the potential integration of LLMs into future critical care workflows.

## Results

Mean age of the patient cohort was 65.9 +/− 16.4 years, mean Charlson comorbidity index was 5.1 +/− 2.9, and mean APACHE score was 22.1 +/− 6.8. The actual in-hospital mortality rate of the cohort was 17%.

### Predictive Performance

Performance metrics for Gemini, Llama, GPT-4, R1 (with and without additional clinical context), and the APACHE II scoring system are summarized in **Table 1**. Results are reported separately for the baseline (no additional context) and context-enhanced conditions. The APACHE II model classified 58.5% of patients as high risk, 40% as intermediate risk, and 1.5% as low risk, with corresponding observed mortality rates of 26.8%, 3.6%, and 0%, respectively. Of all LLMs assessed, only Gemini, when provided with additional clinical context, classified two patients (2.9%) as low risk; notably, one of these patients died. No other LLM assigned any patients to the low-risk category, suggesting a general propensity for risk overestimation.

**Table 1.**
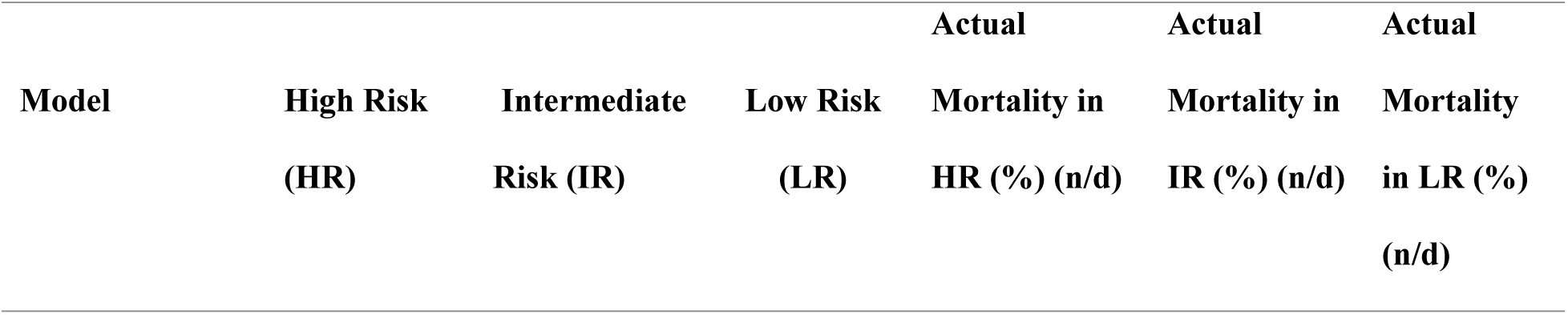
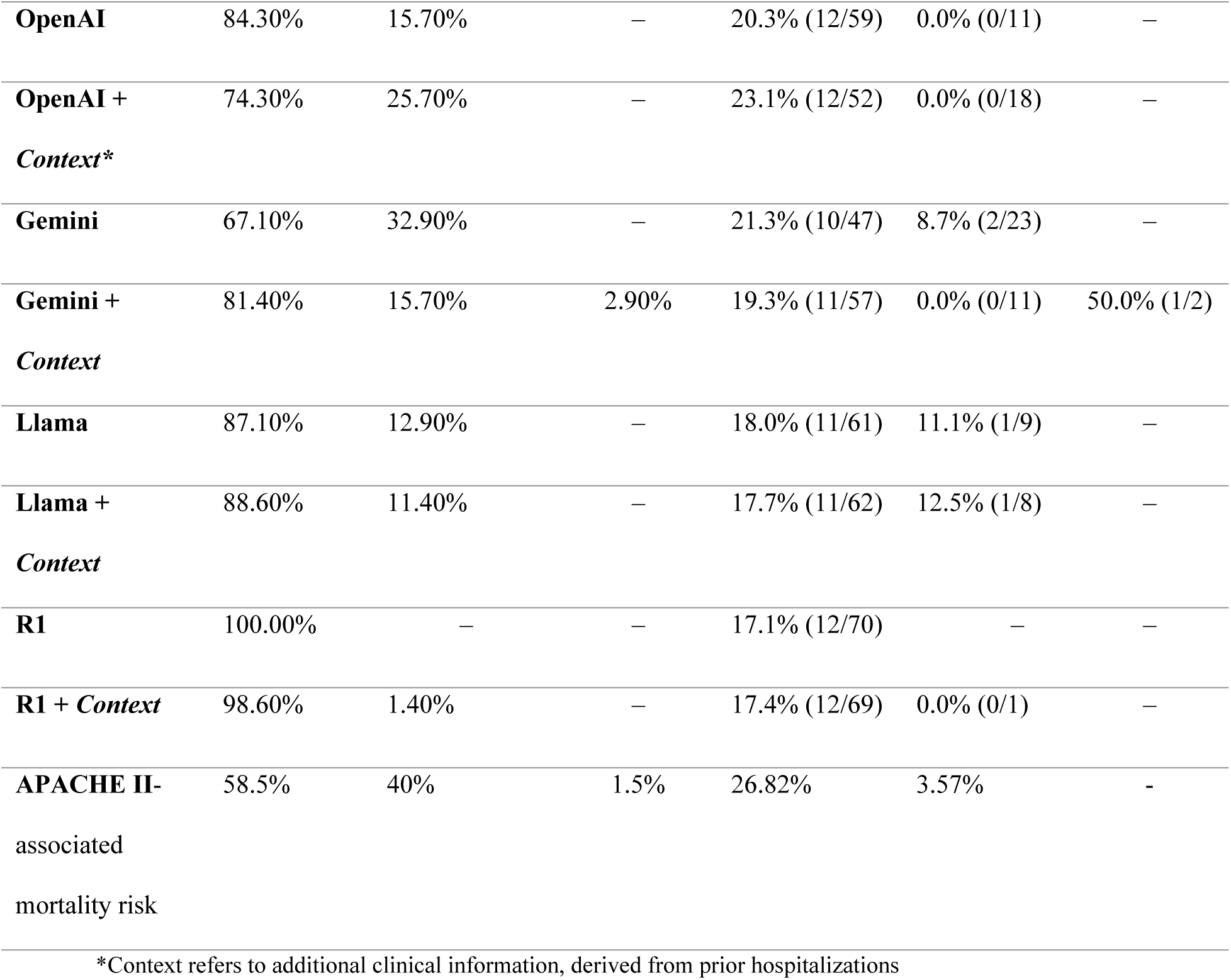
Comparison between the performance of each Large Language Model. The *Context* suffix denotes performance when additional clinical context is provided in the form of prior hospital discharge summaries.

Model calibration, as reflected by observed mortality rates, differed among LLMs, ranging from 17.1% to 23.1%, and was consistently lower than APACHE II estimates, again suggesting a tendency toward over-classification into the high-risk category. Mortality within the intermediate-risk group remained low (0 – 12.5%), consistent with the APACHE II benchmark of 3.57%.

Providing additional clinical context—specifically hospital discharge summaries— impacted both the distribution of mortality risk and model calibration across LLMs. In several cases, the added context led to a more balanced allocation across risk categories. For example, GPT-4 classified fewer patients as high risk when context was included (74.3% with context vs. 84.3% without). Conversely, Gemini increased its high-risk classification with context (81.4% vs. 67.1% without) but was the only model to identify a low-risk group, as noted above. While context altered risk distribution, it did not uniformly improve calibration; in Gemini, high-risk classifications increased after adding context.

### Consistency of Predictions

When evaluated across multiple iterations, Gemini and GPT-4 produced the most consistent predictions (**Fig 1**), suggesting that their probabilistic token generation mechanisms were less susceptible to randomness. The R1 model demonstrated similar performance, though with slightly greater variability than Gemini and GPT-4. Llama exhibited low baseline consistency, which improved after clinical context was incorporated.

**Fig 1.**
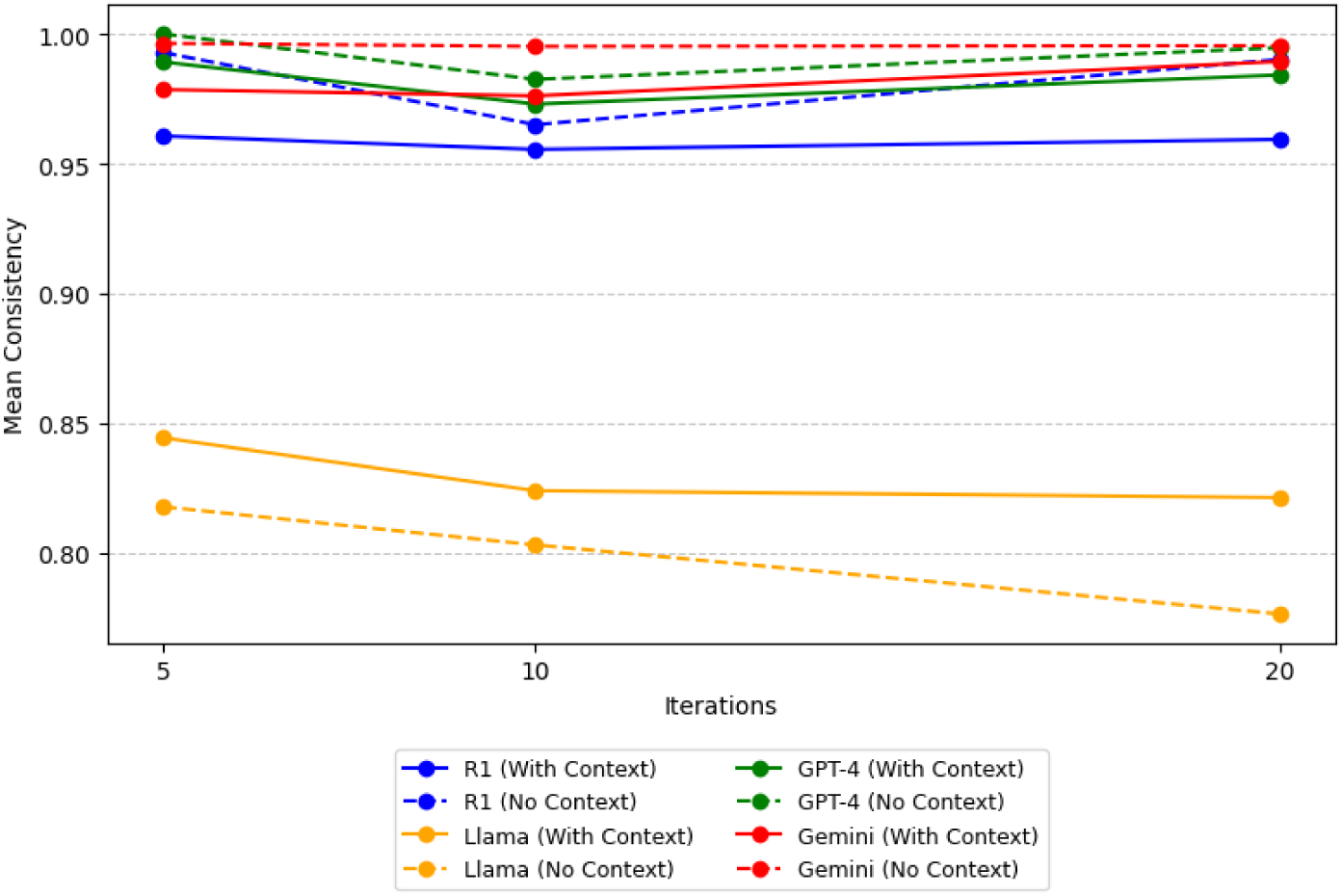
Consistency with respect to First Prognostication Result. The consistency of predictions from various Large Language Models (LLMs)—specifically Gemini, GPT-4, R1, and Llama— varies across multiple iterations (5, 10, and 20 iterations). Consistency scores represent how closely subsequent predictions align with the initial prognostication. Comparisons are made both with and without additional clinical context provided via discharge summaries (from prior hospitalizations). Notably, Gemini and GPT-4 consistently exhibit high consistency scores, indicating stable predictive behavior, irrespective of the presence or absence of additional clinical context. In contrast, Llama shows substantial variability, especially in the absence of clinical context, demonstrating sensitivity to contextual input. The R1 model shows intermediate consistency and demonstrates slight improvement when context is provided.

Analysis of the standard deviation of mortality predictions further supported these findings. Both Gemini and GPT-4, closely followed by R1, exhibited low standard deviations, underscoring their stability and reliability across iterations (**Fig 2**). Interestingly, models provided with additional clinical context exhibited a higher standard deviation than their non-contextual counterparts. A notable exception was Llama, which exhibited the highest standard deviation in the non-contextual analysis, indicating greater variability in its predictions. However, both its consistency and calibration improved with the inclusion of additional clinical context. This finding is particularly noteworthy given that Llama is an open-source model with a relatively small parameter size of 3 billion (B) parameters, highlighting the potential impact of context even in smaller-scale architectures.

**Fig 2.**
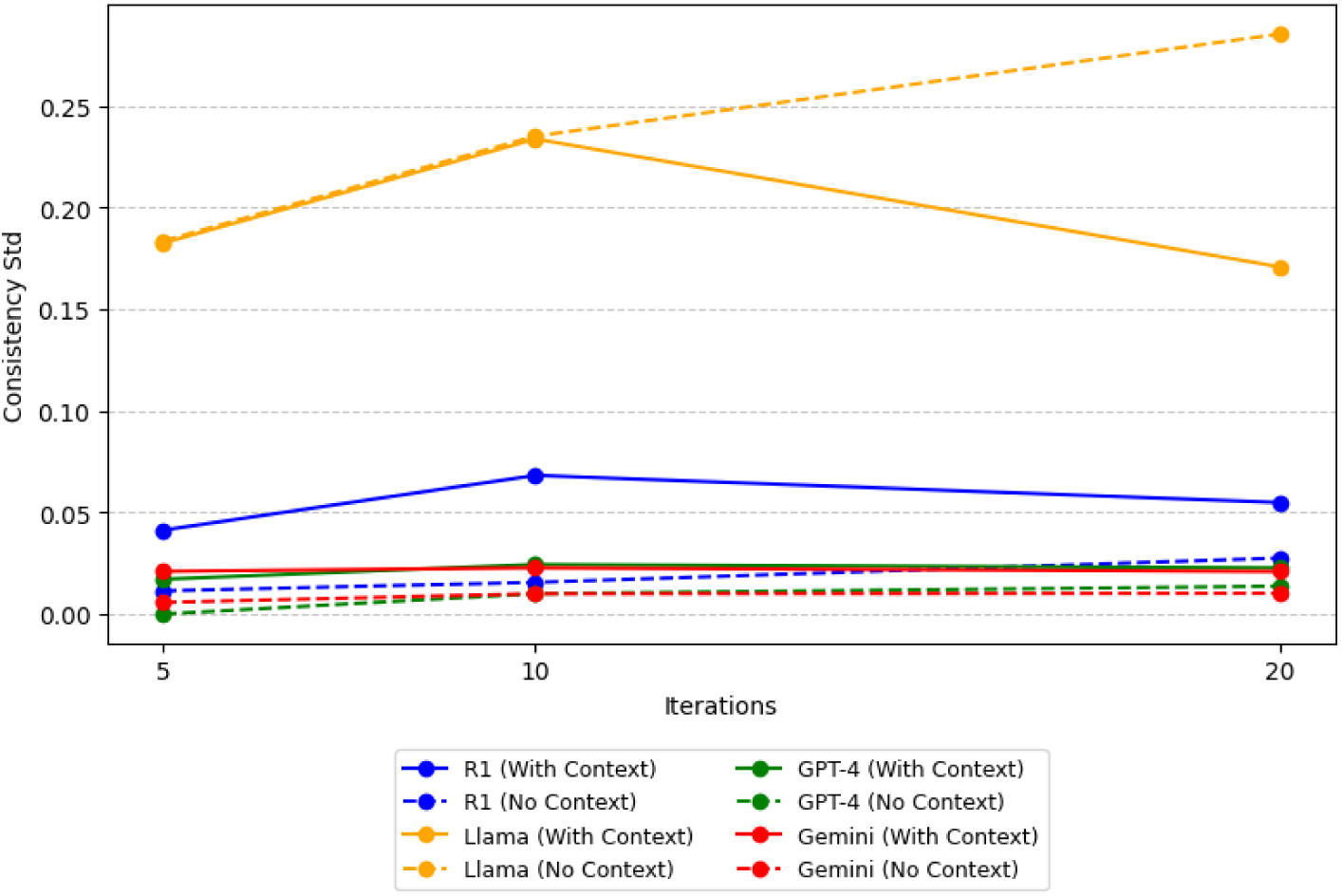
Standard deviation of Prognostication Results. Displayed is the standard deviation of consistency scores from repeated prognostication outputs of the evaluated large language models (LLMs) across increasing iterations (5, 10, and 20). A lower standard deviation indicates greater stability and reliability in model predictions. GPT-4 and Gemini models display notably low standard deviations across all tested conditions, underscoring their robust performance. Conversely, Llama, particularly without added context, exhibits higher variability, reflecting greater uncertainty and less reproducible predictions. The R1 model maintains moderate variability, slightly improving in consistency with additional context, emphasizing the potential benefits of targeted clinical input.

Gemini, GPT-4, and R1 maintained high mean consistency values over multiple iterations (>0.90), potentially making them more reliable for clinical applications. Llama, while potentially more contextually adaptive, suffered from significant inconsistency, which may hinder its utility in scenarios demanding stable outputs. These results underscore the trade-off between consistency and adaptability in LLMs and highlight the need for future work to improve the balance between these characteristics before clinical implementation.

### Rationale for Predictions

**Fig 3** presents the average cosine similarity and standard deviation of rationale statements across multiple iterations for each model. **Supporting Fig S1** presents a representative word cloud comparing differing rationale for risk-stratification following the addition of clinical context. Gemini and GPT-4 consistently exhibited the highest semantic stability, as reflected by their high average cosine similarity and low variability across runs. This indicates that their rationales for assigning patient risk levels remained largely consistent, even when additional clinical context was introduced. These findings suggest that their predictions are guided by internally coherent reasoning rather than stochastic token generation alone.

**Fig 3.**
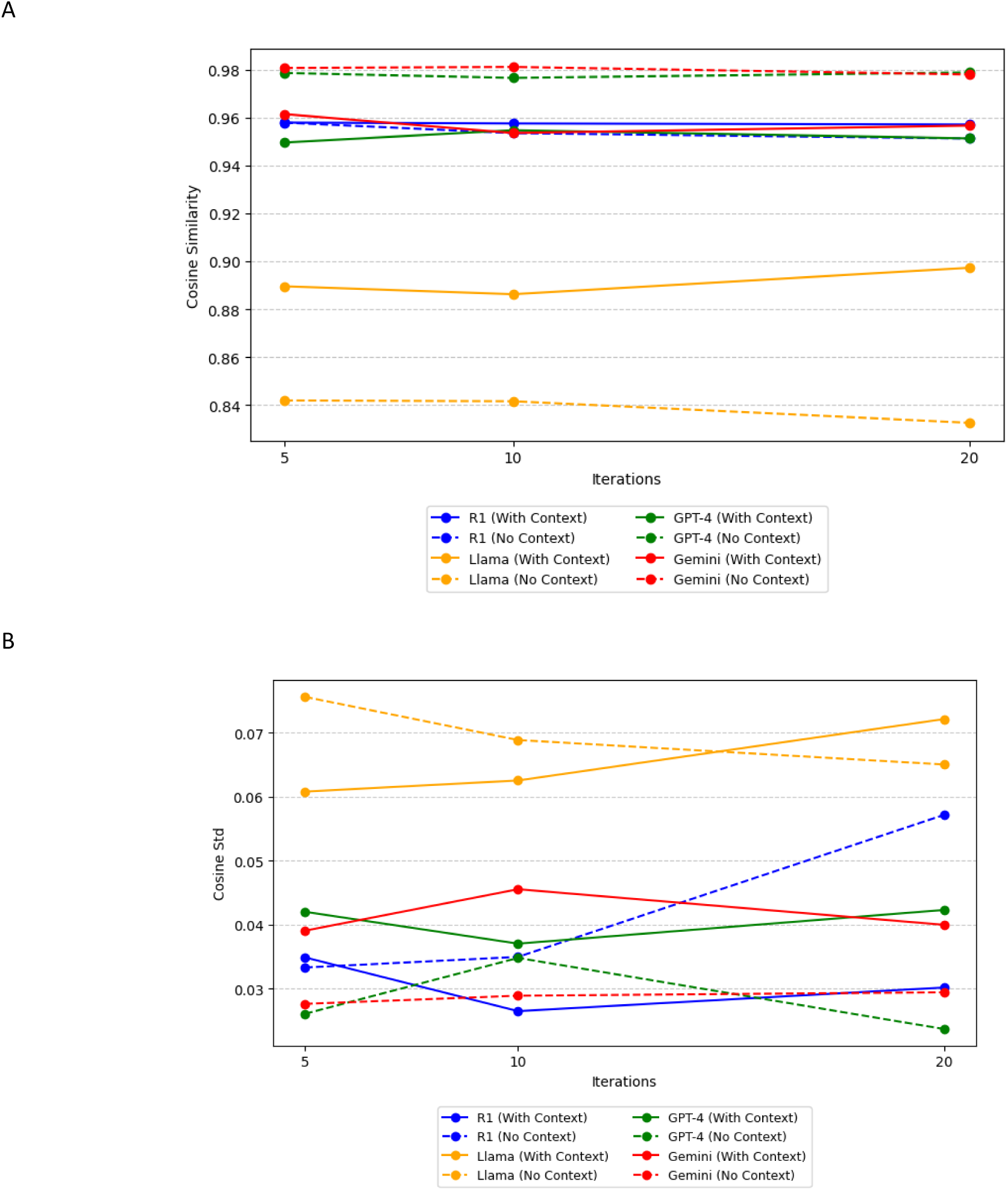
Comparison of the Cosine Similarity Metric of LLM Prognostication. (A) Semantic consistency of prediction rationales, measured by cosine similarity, across repeated iterations. Gemini and GPT-4 maintain high semantic coherence irrespective of added clinical context, whereas R1 and Llama significantly improve with the inclusion of discharge summaries, indicating greater reliance on contextual inputs. (B) Standard deviation of cosine similarity across iterations. Gemini and GPT-4 exhibit low variability, underscoring semantic stability. Llama shows substantial improvement (reduced variability) when provided with clinical context, with R1 demonstrating moderate contextual sensitivity.

In contrast, the Llama model showed the most pronounced improvement in semantic consistency with the addition of clinical context. Its average cosine similarity increased significantly, and variability decreased, suggesting that Llama’s reasoning processes became more stable and interpretable when richer input data were available. This aligns with earlier observations of improved response consistency in Llama under contextualized prompting.

Overall, these results reinforce that consistency in model rationale is not purely stochastic but reflects context-sensitive interpretative shifts. They also highlight that while larger models like GPT-4 and Gemini maintain stability regardless of context, smaller models such as Llama may derive disproportionate benefit from detailed clinical input. This underscores the value of context-aware prompting strategies, particularly when deploying smaller LLMs in clinical reasoning tasks.

## Discussion

Accurate mortality prediction remains a cornerstone of modern intensive care, enabling clinicians to triage and manage critically ill patients more effectively. This study highlights the potential of LLMs as complementary tools to established scoring systems like APACHE II. Beyond predictive accuracy, a key consideration in clinical deployment is data privacy. In this regard, open-source models that can be securely hosted within institutional firewalls may offer advantages. Our findings show that web-based models such as Gemini and GPT-4 achieved superior performance, consistency, and semantic coherence in their predictive rationales, and with more limited data. However, the performance of smaller, open-source models like Llama improved markedly when additional clinical context was provided, underscoring the importance of rich data inputs. These findings align with prior work emphasizing that contextual augmentation enhances machine learning performance in healthcare (4, 6). Ultimately, this study illustrates the trade-offs between model interpretability, consistency, and reasoning depth—factors that must be carefully balanced when integrating LLMs into clinical workflows, particularly in perioperative and critical care environments (12, 14).

A key insight from this study is the emerging promise of deploying open-source language models on lightweight, resource-constrained devices for clinical applications. The performance of the R1 model—which consists of 14B parameters—was notably competitive when compared with significantly larger models like Gemini and GPT-4. Despite its reduced parameter count, R1 demonstrated substantial semantic consistency and prediction reliability, suggesting that model size alone does not dictate clinical utility. This finding is particularly relevant for point-of-care applications, where internet access may be restricted, and local deployment is necessary for compliance with institutional or regulatory privacy standards.

Smaller models like R1 are similarly well-suited for edge computing environments, including mobile health platforms and bedside clinical decision tools. Prior studies have emphasized the growing viability of parameter-efficient models for clinical natural language processing tasks (15), particularly when fine-tuned with domain-specific data or enhanced through retrieval-augmented generation (16). Their lower computational overhead allows for real-time inference without reliance on GPU clusters or high-bandwidth cloud infrastructure, making them more accessible in resource-limited settings.

This study has several limitations. First, the analysis was conducted using a relatively small sample size of 70 ICU admissions from our Critical Illness Data Biorepository. While sufficient for preliminary comparisons, a larger dataset would provide greater statistical power and allow for more robust generalizability of findings. Prior research has demonstrated the value of larger datasets in validating predictive models and in reducing overfitting, particularly in healthcare AI applications (17). LLM predictions were also evaluated using only retrospective data, which may not fully account for real-time clinical nuances or dynamic patient trajectories. Prospective validation in live clinical environments is necessary to establish practical utility. In this respect, high-performing but open-source LLMs such as R1 should be favored.

Another key limitation of this study is its reliance on de-identified clinical data. While essential for preserving patient privacy, de-identification can strip away important contextual and temporal features that may be critical for accurate prognostication. In parallel, the inherent variability in LLM outputs raises concerns regarding reproducibility in clinical environments. Prompt design, temperature settings, and underlying training biases can significantly influence model behavior, complicating efforts to standardize their use (18). Additionally, the study evaluated only four LLMs alongside the APACHE II scoring system—important benchmarks, but only a subset of the broader landscape of predictive tools. Future investigations should incorporate a wider array of machine learning and statistical models to fully assess the comparative performance of LLMs. Importantly, ethical considerations—including data privacy, algorithmic bias, and the interpretability of AI-generated predictions—must be rigorously addressed before these technologies can be responsibly implemented in critical care settings (19, 20).

## Conclusions

As LLMs continue to evolve, their applications in healthcare grow more diverse, opening new frontiers in patient care, medical education, and data analysis. However, these advancements also raise essential questions about transparency (7), ethical data use, privacy safeguards, and the limits of their capabilities—underscoring the need for ongoing scrutiny and research. Future research should focus on optimizing the calibration of LLMs for specific medical datasets, on improving consistency without sacrificing adaptability, and on integrating these models into clinical workflows in ways that complement existing tools like the APACHE score. Nonetheless, our study demonstrates that integrating context-aware prompts can improve the performance of clinical prognostication models.

## Methods

### Clinical Data

Clinical data were obtained from an institutional Critical Illness Data Biorepository (IRB No. 27129) at a quaternary care, academic medical center, and managed using REDCap (Vanderbilt University, Nashville, TN)—a secure, HIPAA-compliant, metadata-driven, browser-based electronic data capture platform. This database contains information from hundreds of patients with critical illness secondary to sepsis or other non-sepsis etiologies. We selected 70 cases to approximate a normal distribution of APACHE II scores (ranging from 9 to 39), drawn from critically ill patients admitted between March 2021 and February 2024. The in-hospital mortality of each patient was known (ground truth), allowing direct comparison between prognostic predictions generated by the APACHE II score and those generated by each LLM.

Due to the sensitive nature of the data and to ensure patient privacy and regulatory compliance, all clinical data exported from REDCap underwent a multi-layered de-identification process before analysis. First, direct identifiers—including names, medical record numbers, and dates of birth—were removed. Next, quasi-identifiers such as admission and discharge dates, geographic information below the state level, and facility-specific identifiers were generalized or replaced with randomized codes. Two investigators manually and independently verified automated de-identification methods to ensure no sensitive data was inadvertently missed. Free-text fields were carefully reviewed and scrubbed to eliminate any embedded identifiers or narrative content that could inadvertently disclose personal information. Additionally, each participant was assigned a unique study ID to allow longitudinal linkage without revealing identity. This de-identification process was guided by the HIPAA Safe Harbor method and reinforced with institutional data governance standards to ensure the dataset was appropriately anonymized for research use. Open-source models were run on password-protected local workstations.

### Large Language Models

We utilized four LLMs for mortality prediction: Llama 3.2 3B (Meta Platforms Inc., Menlo Park, CA), Gemini 1.5 Flash (Google LLC, Mountain View, CA)(18), DeepSeek R1(21), and GPT-4 (OpenAI L.P., San Francisco, CA)(22). Llama 3.2 is available in different configurations with 1B and 3B parameters and supports a context window of up to 128,000 tokens. The R1 model family, including its distilled variants, is based on architectures from Qwen 2.5 and Llama 3. It comprises models with 1.5B, 7B, 8B, 14B, 32B, and 70B parameters. Despite architectural similarities to Llama 3, the R1 model underwent a distinct reinforcement-learning-based training process, warranting separate evaluation. Gemini 1.5 Flash, accessible via an application programming interface, offers a multimodal capability optimized for speed and efficiency, supporting a context window of up to one million tokens and processing text, images, audio, and video inputs. GPT-4, known for generating human-like text, provides options for an 8,192-token or an extended 32,768-token context window, enabling detailed and complex input handling. In this study we set LLM temperatures to be 0.3.

### Predictive Performance

The APACHE II score stratifies patients into distinct risk categories using a point-based system, assigning scores to clinical variables such as age, blood pressure, respiratory rate, and electrolyte concentrations, among others. For the purposes of this study, and supported by prior literature, patients having APACHE II scores of ≤10 (associated with a predicted mortality rate of under 10%) were considered at “low risk” for mortality (23, 24), those with scores ranging between 10 - 20 (10–30% mortality rate) were at “intermediate risk” (23–25), and those having scores >20 (mortality risk > 30%) were at “high risk” (23, 25, 26).

Based on this framework, each LLM was provided with three hypothetical scenarios, each corresponding to a high, intermediate and low risk patient (by APACHE II score) with critical illness (**S1 Appendix**). However, the models were not instructed to apply the APACHE II method for mortality prediction. Instead, each LLM was allowed to use its own ‘judgment’ to independently stratify patients according to their perceived risk of death. Each LLM was then asked to predict whether each of the 70 patients surveyed fell into high, intermediate or low risk of in-hospital death (**S2 Appendix**). To ensure a standardized comparison of model performance, we analyzed only the initial prediction from each model during the first iteration of a five-round evaluation process.

To further assist in prognostication, we then provided each LLM with additional clinical context in the form of de-identified hospital discharge data. Prognostic analyses were then repeated to evaluate whether the inclusion of this additional clinical context would impact each LLM’s prediction.

### Consistency of Predictions

LLM models operate by calculating the most probable next token, a powerful technique that comes with a notable limitation: inconsistency. The same prompt can yield different responses. We evaluate the consistency of the models by repeatedly prompting them with identical inputs over 5, 10, and 20 iterations. The varying number of iterations allowed us to assess the convergence of these models in a medical context. The full dataset of 70 patients was input into each LLM iteratively—first five times, then ten, and finally twenty times. For each patient, the model’s predictions of in-hospital mortality risk were compared across these 5, 10, and 20 iterations to calculate consistency and standard deviation. The consistency score was calculated by comparing the first answer with the answer of each iteration using the following formula:

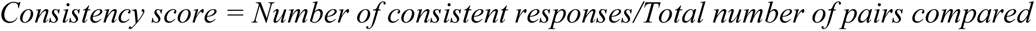

A consistency value closer to 1 indicated less variation in the model’s predictions when provided with identical input data.

### Rationale for Predictions

To determine whether inconsistency in patient risk stratification was purely stochastic or driven by identifiable factors, we analyzed the stated rationales provided by each LLM for categorizing patients as high, intermediate, or low risk. Each rationale was mapped to a medical category using a predefined dictionary, and the cosine similarity between each subsequent iteration and the initial response was calculated. We utilized the SpaCy library (version 3.7.4) along with the pre-trained medium-sized model, *en_core_web_md*, to generate text embeddings. Cosine similarity was then computed between embeddings to quantitatively assess the semantic alignment across iterations.

## Data Availability

The authors will provide the data to those making reasonable request

## Acknowledgements

This study was facilitated by the Penn State Critical Illness and Sepsis Research Center (CISRC, RRID:SCR_026307) of Pennsylvania State University College of Medicine, via the Office of the Vice Dean of Research and Graduate Students. The content is solely the responsibility of the authors and does not necessarily represent the official views of the University or College of Medicine.

## Ethical Considerations

Informed consent was not required for this secondary data analysis, as the study qualified for Exemption under institutional review board (IRB) criteria (IRB #27129). The original (parent) study was approved by the IRB in July 2020 (IRB #15328) and obtained informed consent from patients or from their legally authorized representatives. All human research activities were conducted in accordance with institutional policies aligned with the Declaration of Helsinki and adhered to established standards for data privacy and security.

